# DualStream-MTCA: A Hybrid Deep Learning Architecture for Joint Prediction and Early Detection of Sepsis and Heart Failure in Adult Intensive Care

**DOI:** 10.64898/2026.09.10.26362699

**Authors:** Sako Aras Khdir, Omed Hassan Ahmed

## Abstract

**Objective:** Sepsis and heart failure produce overlapping early physiological signs in intensive care but call for opposite treatments, yet existing models predict one condition at a time. We built one model for both.

**Methods:** DualStream-MTCA combines two bidirectional long short-term memory encoders, for 48 hours of hourly vital signs and 7 days of daily laboratory results, with multi-head cross-attention, gradient-boosted-tree leaf embeddings and two task heads trained under uncertainty weighting. A refined configuration adds per-window aggregates, static patient features, self-supervised encoder pretraining and a five-seed ensemble with Monte Carlo dropout. Development used 53,229 adult stays from MIMIC-IV version 3.1; the frozen models were applied to 102,695 stays from the eICU Collaborative Research Database.

**Results:** At a prediction time 48 hours after admission, the refined ensemble reached an area under the receiver operating characteristic curve of 0.867 for sepsis and 0.899 for heart failure internally, against 0.832 and 0.857 for the baseline model. Isotonic regression brought expected calibration error below 0.015. For positives whose onset followed the prediction time by 4 to 12 hours, the area dropped to 0.66–0.70. Externally, the refined ensemble scored 0.750 for sepsis and 0.786 for heart failure, against 0.711 and 0.821 for the baseline model.

**Conclusion:** A single multi-task model matched its single-task variants on both conditions, but discrimination fell under strictly anticipatory prediction and across sites, where static features carried little usable signal.

**Significance:** To our knowledge, this is the first model built specifically to predict both conditions jointly, with calibration, decision-curve and external evaluation.

## I. Introduction

Sepsis-3 defines sepsis as life-threatening organ dysfunction caused by a dysregulated host response to infection [1]. An estimated 166 million cases and 21.4 million related deaths occur each year [2]. Heart failure affects more than 64 million people, and five-year mortality after symptom onset approaches 50% [3]. Both conditions are common in adult intensive care units (ICUs), and in both, delay is costly. Each hour of antibiotic delay in sepsis is associated with higher mortality, at a pooled odds ratio of 1.04 per hour [4], [5]; heart failure recognized late lengthens ICU stays and pushes patients toward cardiogenic shock [6].

Early on, the two are hard to tell apart. Tachycardia, hypotension, tachypnea and deranged blood chemistry appear in both, and many ICU patients are already abnormal at baseline. The treatments pull in opposite directions: fluid resuscitation and antibiotics for sepsis, diuresis and afterload reduction for decompensated heart failure. Fluid given to the wrong patient risks pulmonary edema; diuresis in a volume-depleted septic patient can precipitate circulatory collapse [7]. Bedside scores such as the Sequential Organ Failure Assessment (SOFA) register organ dysfunction after the fact rather than anticipating it [8], and natriuretic peptides are usually measured only once symptoms appear.

Machine learning on electronic health record (EHR) time series now outperforms conventional screening for sepsis and, increasingly, for heart failure [9], [10]. Nearly all of that work handles one condition at a time. Two independent models give two scores never trained against each other, whereas the bedside question is which process, or both, explains the picture. The nearest published studies predict 28-day mortality among patients already carrying both diagnoses [7], or pick out sepsis-associated acute heart failure inside a septic population [11]. Neither predicts the two conditions jointly from one record.

This paper presents DualStream-MTCA (Dual-Stream Multi-Task Cross-Attention), which predicts both conditions from a single ICU record. We make three contributions. The first is the architecture itself: hourly vital signs and daily laboratory results are encoded in separate bidirectional long short-term memory (BiLSTM) streams, coupled by cross-attention, given gradient-boosted-tree structure through learnable leaf embeddings, and balanced across the two tasks by learned uncertainty. The second is the evaluation on 53,229 MIMIC-IV stays, which covers classical baselines, a seven-variant ablation, and calibration, decision-curve, subgroup, attribution and pre-onset analyses. The third is external: the frozen models are applied to 102,695 stays from the eICU Collaborative Research Database (eICU-CRD), which reveals a transfer trade-off in the static patient features. Negative results are reported as such, among them the components that fail bootstrap tests and the drop in discrimination under strictly anticipatory prediction.

## II. Related Work

Work on sepsis prediction has shifted from tree ensembles and interpretable models over tabular features [9], [12] toward recurrent and attention-based sequence models. DeepAISE joined recurrent encoding to survival modeling [13], Lauritsen *et al*. brought deep learning to EHR event sequences [14], and transformer encoders have served for hours-ahead prediction [15]. Moor *et al*. reported site-dependent AUROCs between 0.75 and 0.85 for models transferred across international cohorts [16]. That pattern matches meta-analytic evidence that external performance usually falls [17], and the degradation observed in a widely deployed proprietary sepsis model [18]. Recent work in this journal combined rule-based explainable models with MIMIC-IV development and eICU-CRD assessment [19]. Heart-failure prediction from EHR data has taken a similar route, with deep models for decompensation and mortality [6], [20] and for risk from short records [21], though gradient-boosted trees remain competitive with cross-attention transformers for early heart-failure prediction [22].

Hard parameter sharing lets related tasks learn a common representation [23], while homoscedastic-uncertainty weighting learns the balance between task losses rather than having it set by hand [24]. Treating the pattern of missing measurements as signal in its own right improves recurrent models on EHR data [25]. Clinical benchmarks do already predict many conditions simultaneously, septicemia and heart failure among 25 code-based whole-stay phenotypes [26], but those labels carry no onset time and neither condition is assessed for calibration or transfer. We are aware of no prior work that builds a joint model dedicated to sepsis and heart failure; the adjacent studies [7], [11] condition one diagnosis on the other.

## III. Methods

### A. Data and Cohort

We used MIMIC-IV v3.1 [27], [28] via PhysioNet [29]: 94,458 ICU stays from a single academic medical center, 2008– 2022. Six filters were applied in order: age of at least 18 years; ICU length of stay of at least 24 h; no do-not-resuscitate or do-not-intubate order at admission; valid identifiers; first eligible stay per patient; and at least one vital-sign record in the first 24 h. What remained was 53,229 stays. Since each patient contributes a single stay, a 70/15/15 split stratified jointly on the two labels is patient-disjoint; it yields 37,260 training, 7,984 validation and 7,985 test stays. Model selection, calibration fitting and early stopping used the validation split; each reported model was scored once on the test split.

### B. Outcome Labels

Sepsis labels followed the Sepsis-3 operationalization of Seymour *et al*. [8]. Suspected infection required either an antibiotic followed by a culture within 72 h or a culture followed by an antibiotic within 24 h, and the earlier of the two events defines *t*_susp_. Hourly SOFA was computed across six organ systems, with two simplifications that are common in the literature: the cardiovascular component uses mean arterial pressure (MAP) below 70 mmHg together with any vasopressor and ignores dose tiers, and the renal component rests on creatinine alone. With *W* = [*t*_susp_ − 48 h, *t*_susp_ + 24 h] and ΔSOFA(*t*) = SOFA(*t*) − SOFA(*t*_susp_ − 48 h), sepsis onset is

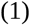

and a stay with no qualifying hour is negative. A stay was labeled positive for heart failure if it had an ICD-10 code beginning with I50, or any N-terminal pro-B-type natriuretic peptide (NT-proBNP) measurement above 400 pg/mL. Each source misses cases for its own reason: a code is absent when heart failure is managed but never coded, and NT-proBNP is absent when nobody ordered the test. Onset is the first qualifying NT-proBNP measurement, or admission where only the code applies. Sepsis prevalence was 52.65% and heart-failure prevalence 25.83%, with 15.70% of stays positive for both. The high sepsis figure is a consequence of strict Sepsis-3 labeling; studies that label from discharge codes usually report 10–25%.

### C. Features and Prediction Time

The prediction time was fixed for every stay at *t*_pred_ = ICU admission + 48 h. The vitals stream comprises seven hourly signals (heart rate, systolic, diastolic and mean arterial pressure, respiratory rate, oxygen saturation and temperature) over the 48 h preceding *t*_pred_, averaged within each hour and forward-filled for up to 4 h. The labs stream holds 15 daily values (white-cell count, lactate, NT-proBNP, creatinine, troponin T, hemoglobin, platelets, bilirubin, alanine and aspartate aminotransferase, albumin, glucose, potassium, sodium and blood urea nitrogen) over the 7 days preceding *t*_pred_, forward-filled for up to 48 h. Implausible values were treated as missing. A binary missingness mask is concatenated to each stream, which makes the tensors 48 × 14 and 7 × 30. Gaps that remained were filled with training-split medians, and all features were z-scored using training-split statistics. The latest value of each laboratory feature, **x**_latest_ ∈ ℝ^15^, is held separately for the tree branch. Onset precedes *t*_pred_ in most positives, so the headline metrics measure detection at a fixed 48-h snapshot; pre-onset performance is reported separately (Section III-H).

### D. Architecture

Fig. 1 lays out the model. A two-layer BiLSTM [30] with 128 units per direction encodes each stream, producing **H**^(v)^ ∈ ℝ^48×256^ and **H**^(l)^ ∈ ℝ^7×256^. An eight-head cross-attention block [31] then lets each stream query the other, taking queries from one stream and keys and values from its counterpart:

**Fig. 1.**
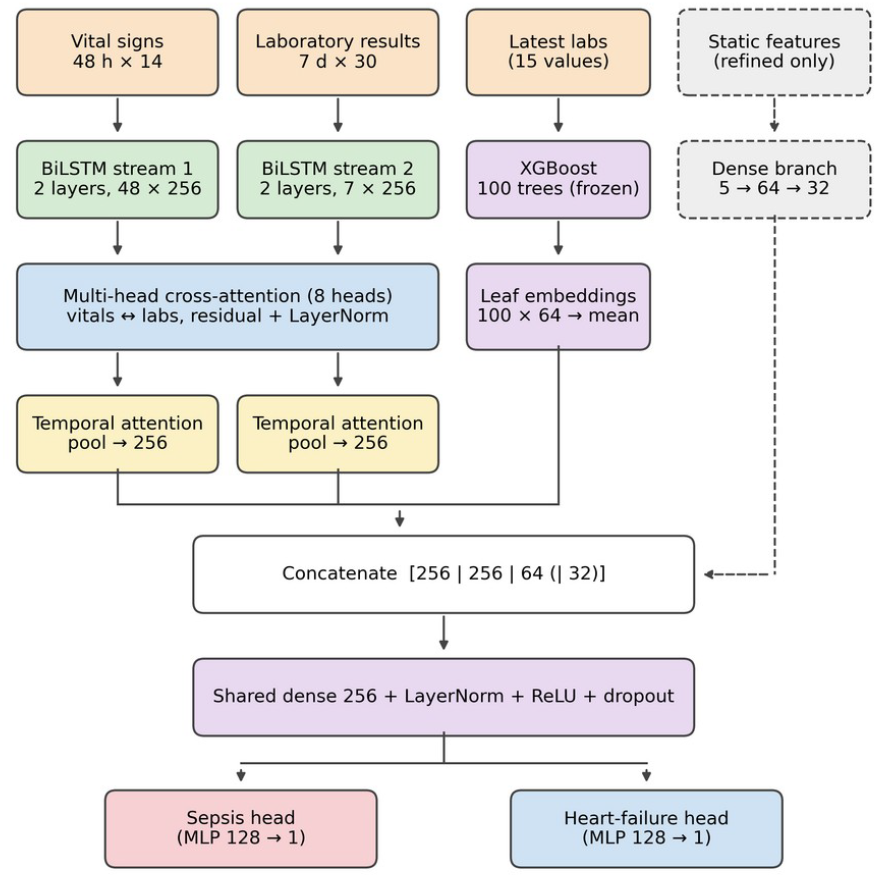
DualStream-MTCA architecture. Dashed elements belong to the refined configuration only.

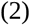

with *d*_k_ = 32, wrapped in a residual connection, layer normalization and a feed-forward layer. Additive temporal-attention pooling [32] then reduces each attended sequence,

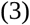

where *s*_t_ is produced by a two-layer scoring network, yielding **z**_v_, **z**_l_ ∈ ℝ^256^. Alongside this, an XGBoost classifier [33] of 100 trees is fitted on **x**_latest_ in the training split against the target sepsis OR heart failure, then frozen. Its 100 leaf indices, one per tree, select learnable 64-dimensional embeddings for each stay; these are mean-pooled into **z**_xgb_ and learned by backpropagation. The concatenation [**z**_v_; **z**_l_; **z**_xgb_] ∈ ℝ^576^ passes through a shared dense layer (256 units, layer normalization, ReLU and dropout) before reaching the two task heads, each a two-layer perceptron with 128 hidden units.

### E. Training Objective

For task *k*, the loss is class-weighted binary cross-entropy,

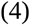

where *w*_k_ ^+^ is the training-split ratio of negatives to positives. Homoscedastic-uncertainty weighting [24] combines the two task losses,

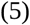

where *σ*_S_ and *σ*_H_ are learned jointly with the network. Optimization used AdamW [34] at learning rate 10^−3^ with weight decay 0.01, cosine annealing after a 5-epoch warm-up, gradient clipping at norm 1.0, batch size 128, mixed precision, and early stopping on validation loss with patience 10. We refer to this configuration as the *baseline model*.

### F. Refined Configuration

The refined configuration retains every component described above and adds four changes. Seven aggregates summarize each time bin: mean, standard deviation, minimum, maximum, slope, last value and count. A static branch (5 → 64 → 32) encodes age, sex, Charlson comorbidity index, number of prior ICU stays and an anchor-year index. Masked reconstruction pretrains both encoders for 30 epochs, minimizing mean squared error on masked timesteps over 74,761 adult MIMIC-IV stays with an ICU length of stay of at least 24 h, a superset of the supervised cohort that includes the input sequences, though not the labels, of the validation and test stays. Last, five models trained from different seeds are averaged, and Monte Carlo dropout [35] over 20 stochastic passes supplies a per-stay uncertainty estimate. We call this the *refined ensemble*.

### G. Comparato

Three classical comparators were trained on the same split from the latest value of the 22 input variables: logistic regression, a 200-tree random forest and a standalone XGBoost classifier. Seven ablations each altered one component of the baseline model: the missingness masks were removed; the two streams were replaced by a single LSTM over concatenated features; the XGBoost branch was removed; mean pooling stood in for attention pooling; cross-attention was removed; and two single-task models were trained, one for sepsis only and one for heart failure only.

### H. Evaluati

We report the area under the receiver operating characteristic curve (AUROC) and the area under the precision–recall curve (AUPRC), each with 95% confidence intervals (CIs) from 1,000 patient-level bootstrap samples. A difference counts as significant only where the CIs fail to overlap, a conservative rule. Calibration was measured by Brier score and expected calibration error (ECE, ten bins). Both Platt scaling [36] and isotonic regression [37] were fitted on the validation split, and whichever gave the lower validation log-loss was applied. Decision-curve analysis [38] served to assess clinical utility. Subgroups by sex, age band and first care unit were scored where they held at least 30 stays and both classes. Feature attributions came from Integrated Gradients [39] over 500 test stays. Pre-onset evaluation retained the test positives whose onset satisfied *t*_onset_ ≥ *t*_pred_ + *h*, for *h* ∈ {4, 6, 8, 12} h, together with all negatives.

For external validation the frozen models were applied, without any retraining, to eICU-CRD v2.0 [40], which spans 208 U.S. hospitals over 2014–2015. Normalization statistics and the XGBoost encoder stayed fixed at their MIMIC-IV values. The same filters applied, apart from the code-status filter, whose source field has no eICU equivalent. Here the SOFA cardiovascular component rested on MAP alone, and missing Glasgow Coma Scale and inspired-oxygen values defaulted to normal. The external cohort held 102,695 stays.

## IV. Results

### A. Internal Discrimination and Calibrati

On the held-out test set the refined ensemble reached an AUROC of 0.867 (95% CI 0.859–0.874) for sepsis and 0.899 (0.892–0.906) for heart failure. The baseline model reached 0.832 and 0.857, and the CIs do not overlap on either task (Table I). The five seeds differed by under 0.005, and Monte Carlo dropout shifted AUROC by less than 0.002 while supplying a mean predictive standard deviation of 0.054 for sepsis and 0.050 for heart failure. Baseline AUPRC was 0.837 for sepsis and 0.720 for heart failure.

**TABLE I.** Internal Test-Set Performance (MIMIC-IV, n = 7,985)

| Metric | Baseline model | Refined ensemble |
| --- | --- | --- |
| Sepsis AUROC | 0.832 (0.824–0.841) | 0.867 (0.859–0.874) |
| HF AUROC | 0.857 (0.848–0.866) | 0.899 (0.892–0.906) |
| Sepsis Brier, raw / cal. | 0.170 / 0.167 | 0.150 / 0.148 |
| HF Brier, raw / cal. | 0.145 / 0.122 | 0.125 / 0.107 |
| Sepsis ECE, raw / cal. | 0.052 / 0.013 | 0.027 / 0.014 |
| HF ECE, raw / cal. | 0.135 / 0.009 | 0.130 / 0.013 |

Parentetical values are bootstrap 95% CIs. Cal., after isotonic regression; HF, heart failure. The refined-ensemble calibration rows were computed without Monte Carlo dropout (AUROC within 0.001); with it, raw ECE is 0.035 for sepsis and 0.127 for HF.

Raw heart-failure probabilities ran overconfident in the upper bins, at a raw ECE of 0.135 for the baseline model and 0.130 for the refined ensemble. Isotonic regression won over Platt scaling on both tasks; it cut ECE to 0.009 for heart failure and from 0.052 to 0.013 for sepsis. In the refined ensemble, ECE fell from 0.027 to 0.014 for sepsis and from 0.130 to 0.013 for heart failure (Fig. 2). Over the clinically plausible range of thresholds, net benefit stayed above both treat-all and treat-none (Fig. 3). At a threshold of 0.20, for instance, heart-failure net benefit for the baseline model was 0.140 against 0.073 for treat-all.

**Fig. 2.**
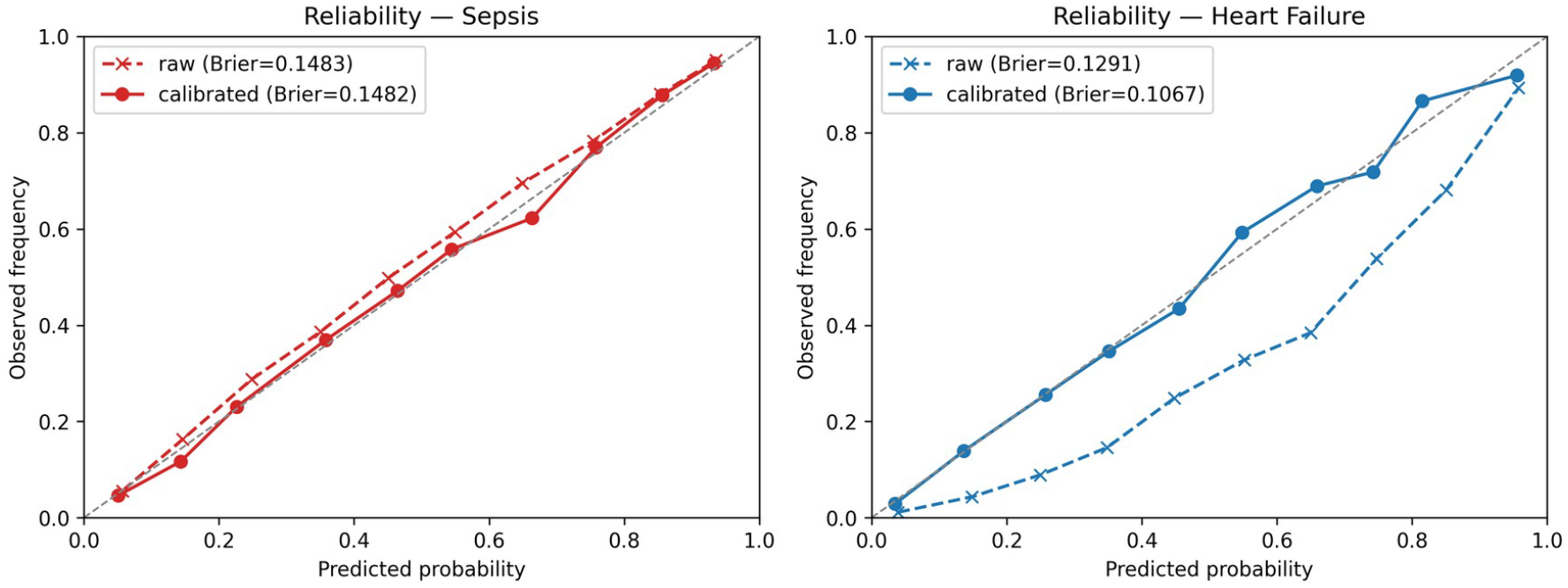
Reliability diagrams for the refined ensemble on the MIMIC-IV test set, shown before (dashed) and after (solid) isotonic calibration, for sepsis (left) and heart failure (right).

**Fig. 3.**
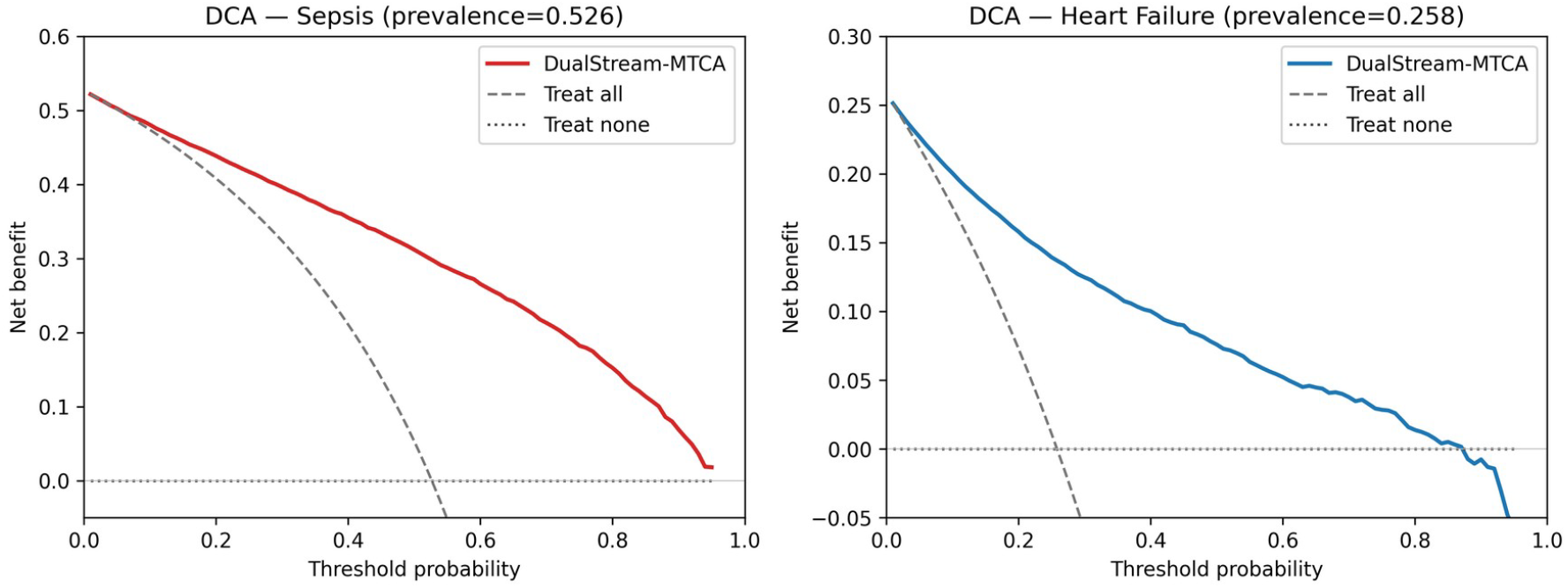
Decision-curve analysis for the refined ensemble on the MIMIC-IV test set, sepsis (left) and heart failure (right), compared against treat-all and treat-none.

### B. Baselines and Ablations

All three classical baselines fell short of the baseline model (Table II). Its sepsis lift over standalone XGBoost, +0.045, was significant; the heart-failure gap of +0.009 sat inside overlapping CIs, consistent with the latest laboratory values, NT-proBNP above all, carrying most of the heart-failure signal. The single-LSTM ablation, the one temporal neural comparator, scored 0.817 for sepsis.

**TABLE II.** Comparison With Baselines (MIMIC-IV Test Set)

| Model | Sepsis AUROC | HF AUROC |
| --- | --- | --- |
| Logistic regression | 0.727 (0.716–0.737) | 0.755 (0.742–0.767) |
| Random forest | 0.784 (0.775–0.794) | 0.845 (0.835–0.855) |
| XGBoost | 0.787 (0.777–0.796) | 0.848 (0.838–0.859) |
| Single LSTM (ablation) | 0.817 | 0.853 |
| Baseline model | 0.832 (0.824–0.841) | 0.857 (0.848–0.866) |

Classical models take the latest value of each of the 22 inputs. Single-LSTM values are the baseline model plus the ablation deltas given in Table III.

Three components proved load-bearing (Table III). Dropping the missingness masks cost 0.018 sepsis AUROC, collapsing the two streams into one LSTM cost 0.015, and dropping the XGBoost branch cost 0.015 heart-failure AUROC. Two changes stayed within bootstrap noise: removing cross-attention, and substituting mean pooling for attention pooling. The single-task models landed inside the CIs of the multi-task model, so the second prediction came at no measurable cost.

**TABLE III.** Ablation Deltas Relative to the Baseline Model.

| Variant | ΔSepsis | ΔHF | Significant |
| --- | --- | --- | --- |
| No missingness masks | -0.018 | -0.006 | Sepsis |
| Single LSTM | -0.015 | -0.004 | Sepsis |
| No XGBoost branch | +0.009 | -0.015 | HF |
| Mean pooling | +0.004 | +0.000 | No |
| No cross-attention | -0.003 | +0.003 | No |
| Sepsis only | +0.002 | — | No |
| HF only | — | +0.000 | No |

Significance by non-overlapping bootstrap 95% CIs.

### C. Pre-onset Subsets

Between 57 and 91 test positives, depending on task and horizon, had an onset at least 4 h past the prediction time, some 2–3% of all positives. AUROC on these subsets fell to 0.66– 0.70 on both tasks (Table IV). AUPRC came to 0.055–0.066, roughly 3–7 times the subset prevalence of 0.9–2.4%. Heart-failure results are identical at 4, 6 and 8 h, because no test-set onset fell between 52 and 56 h after admission.

**TABLE IV.** Pre-Onset Subsets, Baseline Model.

| Horizon | Task | N+ | AUROC | AUPRC |
| --- | --- | --- | --- | --- |
| 4 h | Sepsis | 91 | 0.695 | 0.066 |
| 12 h | Sepsis | 74 | 0.696 | 0.055 |
| 4 h | HF | 64 | 0.671 | 0.066 |
| 12 h | HF | 57 | 0.661 | 0.065 |

Negatives number 3,781 for sepsis and 5,922 for HF. Results at 6 and 8 h lie between those shown.

### D. Subgroups and Attributions

Sepsis AUROC for the refined ensemble declined with age, from 0.882 among patients aged 50 or younger to 0.835 above 80. The largest gain over the baseline model appeared in the cardiac-vascular ICU, where sepsis AUROC rose from 0.752 to 0.871. Differences by sex stayed within 0.005 for the refined ensemble. Among laboratory inputs, Integrated Gradients ranked lactate, troponin T and white-cell count highest for sepsis, and NT-proBNP, lactate, troponin T and blood urea nitrogen for heart failure (Fig. 4). Temperature was the most influential vital sign on both tasks.

**Fig. 4.**
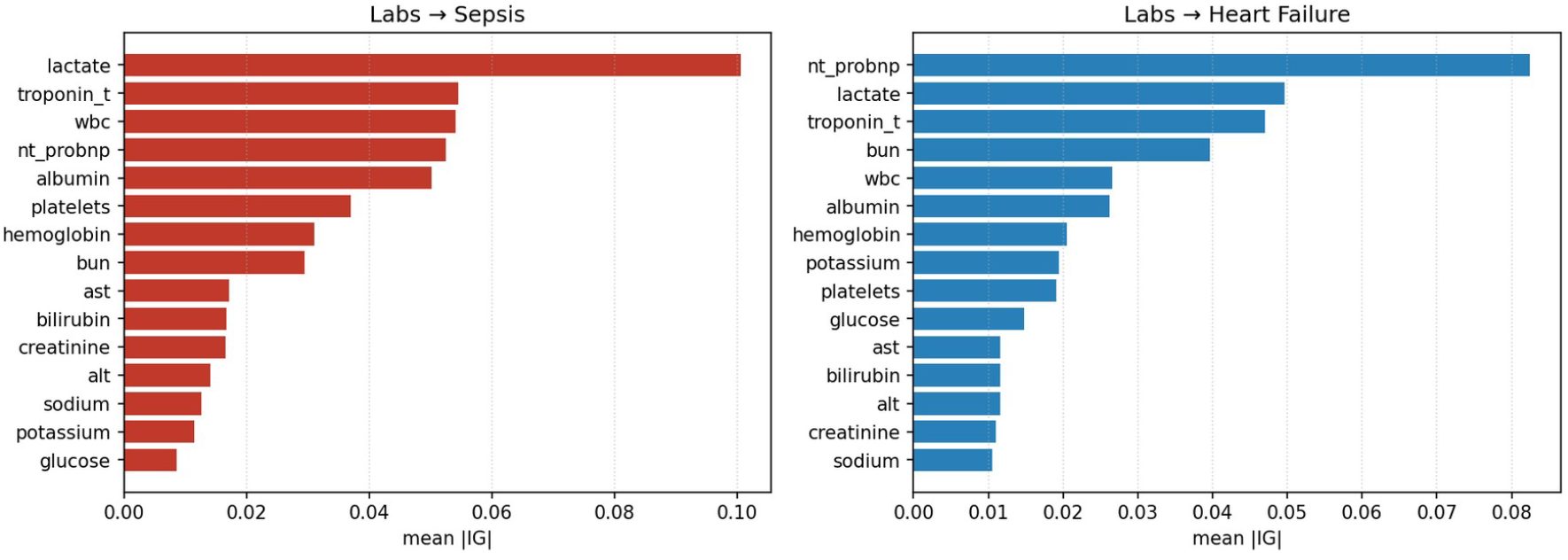
Mean absolute Integrated-Gradients attribution across the laboratory inputs over 500 test stays, for sepsis (left) and heart failure (right).

### E. External Validation

Table V summarizes performance on eICU-CRD. Heart-failure prevalence was 14.19%, while sepsis prevalence was only 0.14%, or 140 of 102,695 stays: just 198 stays met the suspected-infection criterion, because eICU holds roughly 30 times fewer microbiology entries per stay than MIMIC-IV. The baseline model gave up 0.036 heart-failure AUROC, from 0.857 to 0.821, and 0.121 sepsis AUROC, from 0.832 to 0.711. Externally the refined ensemble outperformed the baseline model on sepsis (0.750) but not on heart failure (0.786), a fall of 0.113 from its internal 0.899 (Fig. 5). External sepsis AUPRC was 0.003 against a prevalence of 0.0014.

**TABLE V.** External Validation on Eicu-crd (n = 102,695)

| Task | Baseline model | Refined ensemble |
| --- | --- | --- |
| Sepsis AUROC | 0.711 | 0.750 (0.717–0.781) |
| Sepsis AUPRC | — | 0.003 |
| HF AUROC | 0.821 | 0.786 (0.782–0.790) |
| HF AUPRC | — | 0.428 |
| Internal → external ΔAUROC | −0.121 / −0.036 | −0.117 / −0.113 |

The last row lists sepsis / HF. Prevalence: sepsis 0.14%, HF 14.19%. CIs were not recorded for the baseline model.

**Fig. 5.**
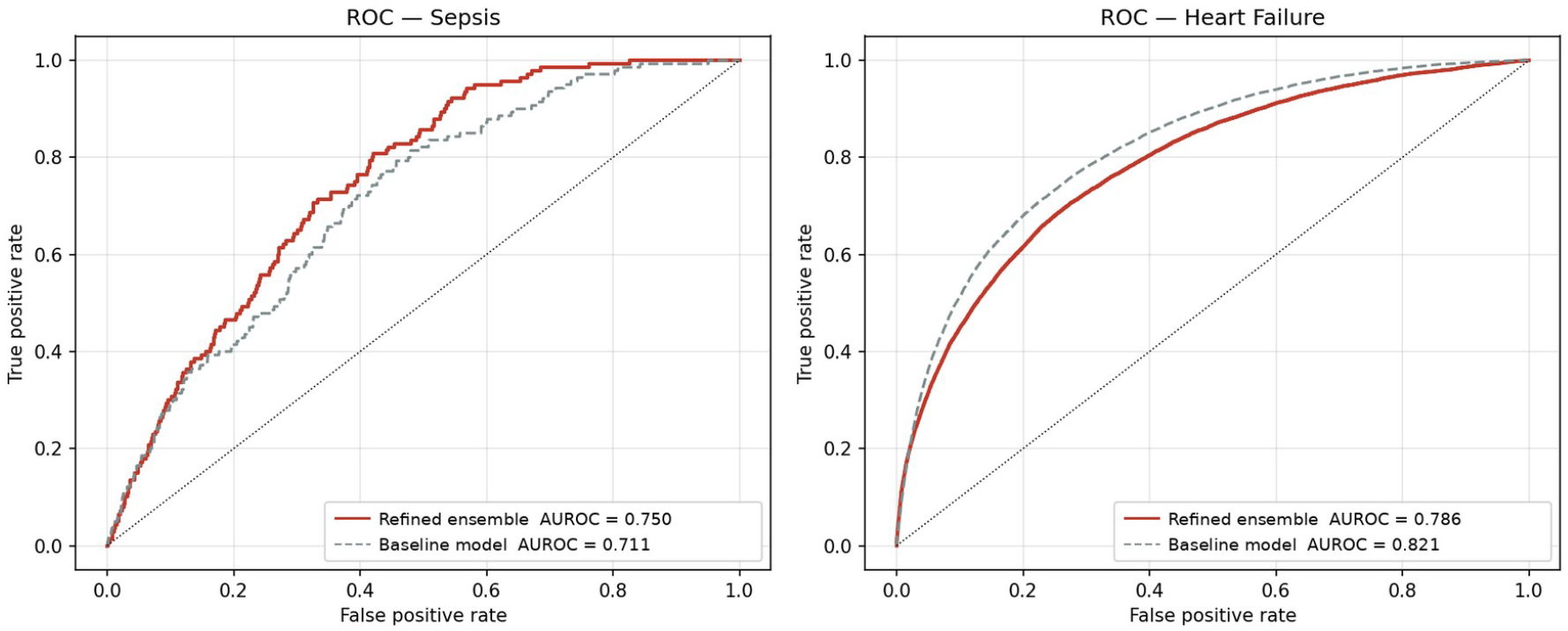
External ROC curves on eICU-CRD for the refined ensemble and for the baseline model, sepsis (left) and heart failure (right).

Much of the heart-failure loss probably traces to the static branch. Disabling it on the internal test set lowered heart-failure AUROC by 0.029, with non-overlapping CIs, and sepsis AUROC by 0.002. That configuration was never evaluated on eICU-CRD, so its external effect remains untested. On eICU the mean Charlson index was 0.06 and the anchor-year index took a single value, leaving neither feature with usable signal. Applying the MIMIC-IV isotonic mapping lowered external heart-failure ECE from 0.181 to 0.058, but raised sepsis ECE from 0.330 to 0.370.

## V. Discussion

One multi-task model predicted both conditions as well as its single-task counterparts did, so a deployment can cover both for the cost of one. Three design choices stand out in the ablations: explicit missingness masks, separate encoders for vitals and laboratory results, and tree-based leaf embeddings over the latest laboratory values. What produces the dual-stream gain is the separation of the streams, not the cross-attention; once the streams are already separate, removing cross-attention moved AUROC by less than 0.005. We keep the block because it is the only point where the streams exchange information before pooling, though its benefit here is unproven.

Internally the refinement stack added 0.035 sepsis and 0.042 heart-failure AUROC, though not all of that gain survives transfer. The static features encode priors specific to MIMIC-IV, so a new site should refit or disable the static branch and refit calibration locally [41]. On heart failure the model sits close to what XGBoost achieves from the latest laboratory values alone, which fits the observation that NT-proBNP is at once the strongest input and part of the label.

### A. Limitations

First, the headline performance is not pre-onset prediction. The prediction time sits 48 h after admission, and onset precedes it in roughly 97–98% of positives, so the headline AUROCs largely measure recognition of established or evolving disease. The strictly anticipatory figures, 0.66–0.70, rest on 57–91 positives, and those subsets were built by filtering the 48-h predictions rather than by re-extracting features at onset minus *h*. Second, labels and inputs overlap. The heart-failure label takes in NT-proBNP above 400 pg/mL, while NT-proBNP is itself an input and the top attribution; the SOFA-based sepsis label shares MAP, platelets, bilirubin and creatinine with the inputs. A sensitivity analysis that drops these inputs, or that ends the windows before onset, is still needed. Third, the sepsis prevalence of 52.65% follows from strict Sepsis-3 labeling with simplified SOFA components, and the heart-failure label marks a disease state rather than acute decompensation. Fourth, the comparators leave out recent temporal architectures: gated recurrent units, temporal convolutional networks and transformers [15]. Fifth, self-supervised pretraining saw the unlabeled inputs of every validation and test stay, so some part of the refined model’s internal gain over the baseline may come from transductive exposure to the test inputs, the baseline model having no pretraining stage at all. Sixth, external sepsis labels are unreliable in eICU-CRD, which means the external sepsis result mainly reflects label ascertainment; higher external sepsis AUROCs have been reported on eICU-CRD under other cohort and label definitions [19]. Both cohorts are from the United States. Finally, evaluation is retrospective; prospective studies must assess effects on treatment timing and alert burden [42].

## VI. Conclusion

DualStream-MTCA predicts sepsis and heart failure jointly from routine ICU data, matches its single-task variants, calibrates well after isotonic regression and offers net benefit over default strategies at a fixed 48-h prediction time. Discrimination drops for strictly pre-onset prediction and across sites, where static features transfer poorly. Pre-onset feature windows, label-independent inputs and modern temporal baselines are the next steps before prospective evaluation.

## Data Availability

The MIMIC-IV and eICU-CRD datasets are available via PhysioNet subject to credentialed access. Code for the preprocessing pipeline, architecture, and evaluation is available upon reasonable request from the corresponding author.

https://physionet.org/content/mimiciv/

https://physionet.org/content/eicu-crd/

## Acknowledgment

Generative AI tools (Claude, Anthropic; Gemini, Google) were used in preparing this manuscript: to restructure and revise the text of all sections, to reformat it to IEEE style, to edit language and grammar, to redraw Fig. 1 from the authors’ original design, to prepare the graphical abstract, and to check reference details. The study design, code, experiments and all reported results are the authors’ own work. The authors reviewed and edited all AI-assisted content and take full responsibility for the manuscript.

## Data and Code Availability

MIMIC-IV and eICU-CRD are available through PhysioNet to credentialed users who complete the required training and sign the data use agreements. Code for preprocessing, the model and evaluation is available from the corresponding author on reasonable request.

## Ethics Statement

This retrospective study used only publicly available, fully de-identified data from MIMIC-IV [27], [28] and eICU-CRD [40], accessed through PhysioNet [29] under credentialed data use agreements. Creation of MIMIC-IV was approved by the institutional review boards of Beth Israel Deaconess Medical Center and the Massachusetts Institute of Technology, and eICU-CRD was deemed exempt by the Massachusetts Institute of Technology review board because it is de-identified. No further institutional review board approval or informed consent was required for this secondary analysis.

